# Differentiating COVID-19 from other types of pneumonia with convolutional neural networks

**DOI:** 10.1101/2020.05.26.20113761

**Authors:** Ilker Ozsahin, Confidence Onyebuchi, Boran Sekeroglu

**Affiliations:** Near East University, Nicosia, Mersin-10 TRNC, 99138 Turkey; Cyprus International University, Nicosia, Mersin-10 TRNC, 99258

**Keywords:** COVID-19, coronavirus, pneumonia, X-ray, convolutional neural networks

## Abstract

**INTRODUCTION:** A widely-used method for diagnosing COVID-19 is the nucleic acid test based on real-time reverse transcriptase-polymerase chain reaction (RT-PCR). However, the sensitivity of real time RT-PCR tests is low and it can take up to 8 hours to receive the test results. Radiologic methods can provide higher sensitivity. The aim of this study is to investigate the use of X-ray and convolutional neural networks for the diagnosis of COVID-19 and to differentiate it from viral and/or bacterial pneumonia, as 2-class (bacterial pneumonia vs COVID-19 and viral pneumonia vs COVID-19) and 3- class (bacterial pneumonia, COVID-19, and healthy group (BCH), and among viral pneumonia, COVID- 19, and healthy group (VCH)) experiments.

**METHODS:** 225 COVID-19, 1,583 healthy control, 2,780 bacterial pneumonia, and 1,493 viral pneumonia chest X-ray images were used. 2-class- and 3-class-experiments were performed with different convolutional neural network (ConvNet) architectures, with different variations of convolutional layers and fully-connected layers.

**RESULTS:** The results showed that bacterial pneumonia vs COVID-19 and viral pneumonia vs COVID- 19 reached a mean ROC AUC of 97.32% and 96.80%, respectively. In the 3-class-experiments, macro-average F1 scores of 95.79% and 94.59% were obtained in terms of detecting COVID-19 among BCH and VCH, respectively.

**CONCLUSIONS:** The ConvNet was able to distinguish the COVID-19 images among non-COVID-19 images, namely bacterial and viral pneumonia as well as normal X-ray images.

## Introduction

The novel coronavirus disease (COVID-19) was first discovered in the city of Wuhan in Hubei province, central China. COVID-19 has been classified as a member of the Orthocoronavirinae subfamily under the Corona viridae family [1]. The members of this family are generally categorized as zoonotic viruses and transmission to humans occurs as a result of contact with infected animals. Bats and snakes have been implicated as the major reservoirs of coronavirus [2]. COVID-19, a novel strain of coronavirus, has an 80% similar identity with severe acute respiratory syndrome coronavirus (SARS-CoV) as the strains share similar lipid rafts. Human-to-human transmission through respiratory droplets has been the major infection route and has caused the spread of the virus [3]. The potential for the virus to spread widely resulted in a global pandemic in 2020, which caused the World Health Organization (WHO) to declare a public health emergency of international concern (PHEIC) [4] on 30th January 2020. As of the time of writing (on May 17, 2020), there are approximately 4,500,00 confirmed cases of human COVID-19 infections in 213 countries, areas, or territories with more than 300,000 deaths worldwide [4]. The primary target of the virus is the lower respiratory tract, which subsequently leads to distress or multiple organ failure in severe cases due to replication of the virus [5]. Lack of access to critical early detections and diagnostic tools has contributed to patient death [6]. A widely used method for diagnosing COVID- 19 infections is the nucleic acid test based on real-time reverse transcriptase-polymerase chain reaction (RT-PCR). However, the sensitivity of real time RT-PCR tests is low, which means it is not adequate for diagnosis, isolation, and contact tracing of infected patients [7]. In addition, the process of obtaining the results with real-time RT-PCR test can take between 3 and 8 hours. Due to the aforementioned reasons, there is a need to investigate other timely approaches for the diagnosis of COVID-19. Since its introduction in the 1970s, CT scanning has revolutionized diagnostic decision making and has had a major impact on the field of surgery, where it has decreased the need for surgery from 13% to 5% and has made many exploratory surgical procedures extinct [8]. Several studies have reported high specificity in differentiating COVID-19 from viral pneumonia on chest CT [7, 9]. However, associated radiation exposure incurred by patients is one of the downsides of CT. This assertion is supported by a study conducted in the United States in 2009, which proposed that 75.4% of the effective radiation dose delivered from all imaging procedures of CT, while X-ray based examination accounts for only 11% [8]. X-ray is a non-invasive imaging approach that is widely available in emergency and hospital settings, where the interpretation of results can often be performed without requiring the input of expert radiologists. X-ray imaging has the potential to serve as an easy method of screening and detecting certain manifestations in the lungs associated with COVID-19 disease without the risk of aerosolization of the pathogens due to probing of the patient’s respiratory system, as is the case with laboratory tests. Despite its advantages, the symptoms detected using X-ray imaging could be common to both COVID-19 and other lung diseases making it difficult to differentiate from other types of pneumonia. Therefore, there is a need for computerized support systems for accurate and precise diagnosis.

Radiography examination for COVID-19 diagnosis can be a good complement to PCR [10]. However, one of the greatest bottlenecks faced is the need for an expert radiologist to interpret the radiography images. This procedure is even more difficult in differentiating COVID-19 from other types of pneumonia since the visual indicators can be subtle [9]. Furthermore, CT exams take significantly more time than X-ray imaging.

Recently, researchers have been working on the development of artificial intelligent (AI) systems for improving the accuracy and speed of the process of interpreting the the radiologic images of COVID-19 patients [11-13]. The study conducted by Wang et al. [11] on the application of a deep learning algorithm using CT images to screen for COVID-19 revealed that out of 54 COVID-19 images, 46 were predicted as COVID-19 positive by the algorithm, with an accuracy of 85.2%. This result indicated the significant value of using the deep learning method to extract radiological graphical features for COVID-19 diagnosis.

The study conducted by Butt et al. [14] on the screening of coronavirus disease pneumonia from other types of pneumonia using AI deep learning models and CT scan achieved an accuracy of 86.7% for the classification of COVID 19 and non-COVID-19 cases. In another study by Sethy and Behera [15], the detection of COVID-19 based deep learning models and X-ray images achieved an accuracy of 95.38% using the ResNet plus SVM model.

Since researchers have only recently started working on the use of AI for COVID-19 detection, many studies have focused on binary classification, usually differentiating COVID-19 patients from a healthy group. In this study, we propose the use of X-ray and convolutional neural networks for the diagnosis of COVID-19 in order to differentiate it from viral and/or bacterial pneumonia, as 2-class and 3-class experiments.

## Materials and methods

### Dataset

A total of 225 COVID-19 chest X-ray images were obtained from [16], which can be accessed from the https://github.com/ieee8023/covid-chestxray-dataset. The project built for creating this dataset was approved by the University of Montreal’s Ethics Committee #CERSES-20-058-D. Complete metadata is not provided for the COVID-19 dataset. Based on the provided information, there were 131 male patients and 64 female patients, and the average age for the COVID-19 group was 58.8±14.9 years. 1,583 healthy control, 2,780 bacterial pneumonia, and 1,493 viral pneumonia chest X-ray images were obtained from [17]. This dataset is available at https://doi.org/10.17632/rscbjbr9sj.3. Informed consent was obtained for all subjects, and the study was approved by the relevant institutional review board at each data acquisition site. All methods were performed in accordance with the relevant guidelines and regulations. All chest X-ray imaging was performed as part of patients’ routine clinical care. Institutional Review Board (IRB)/Ethics Committee approvals were obtained. The work was conducted in a manner compliant with the United States Health Insurance Portability and Accountability Act (HIPAA) and was adherent to the tenets of the Declaration of Helsinki.

### Experimental design

Several experiments were performed by considering the different combinations of datasets and they were divided into two main categories. The 2-class-experiments, which include two diseases at a time, consist of several experiments for the classification of bacterial pneumonia vs COVID-19, and viral pneumonia vs COVID-19. On the other hand, the 3-class-experiments consist of the classification of bacterial pneumonia vs COVID-19 vs healthy control (BCH), and viral pneumonia vs COVID-19 vs health control (BVH).

Different convolutional neural network (ConvNet) architectures with different variations of convolutional layers and fully-connected layers were implemented for each experiment in each category to determine the optimal architecture for the classification. Because of the imbalanced nature of the data, the experiments were repeated with augmented data by increasing the number of training images for the class with the minimum number of images, which also provided more effective convergence of ConvNet. Data augmentation is a procedure that creates new data similar to original data when the data is insufficient for the convergence or is imbalanced. Several methods can be applied in a data augmentation procedure to generate artificial images such as rotation, translation, zoom, etc. However, different approaches that produce more similar images may provide more efficient data for neural networks to boost the convergence. In this study, the data augmentation procedure was applied to the images of COVID-19 class, which had the lowest number of images among all the classes. Four basic and common filtering approaches, namely the Gaussian filter (3×3), bilateral filter (15×15), mean filter (5×5) and the median filter (5×5), were applied to the COVID-19 X-ray images, and blurred images were generated. Figure 1 presents the original image and the generated images in the data augmentation procedure. The use of augmented data in the testing of the images artificially increased the accuracy of the system, while the similar images were sent to the model during the training phase, even if the test images were not used in the training. Therefore, generated data should not be considered in the test phase to assess the real performance of a model on real-life images.

**Figure 1.**
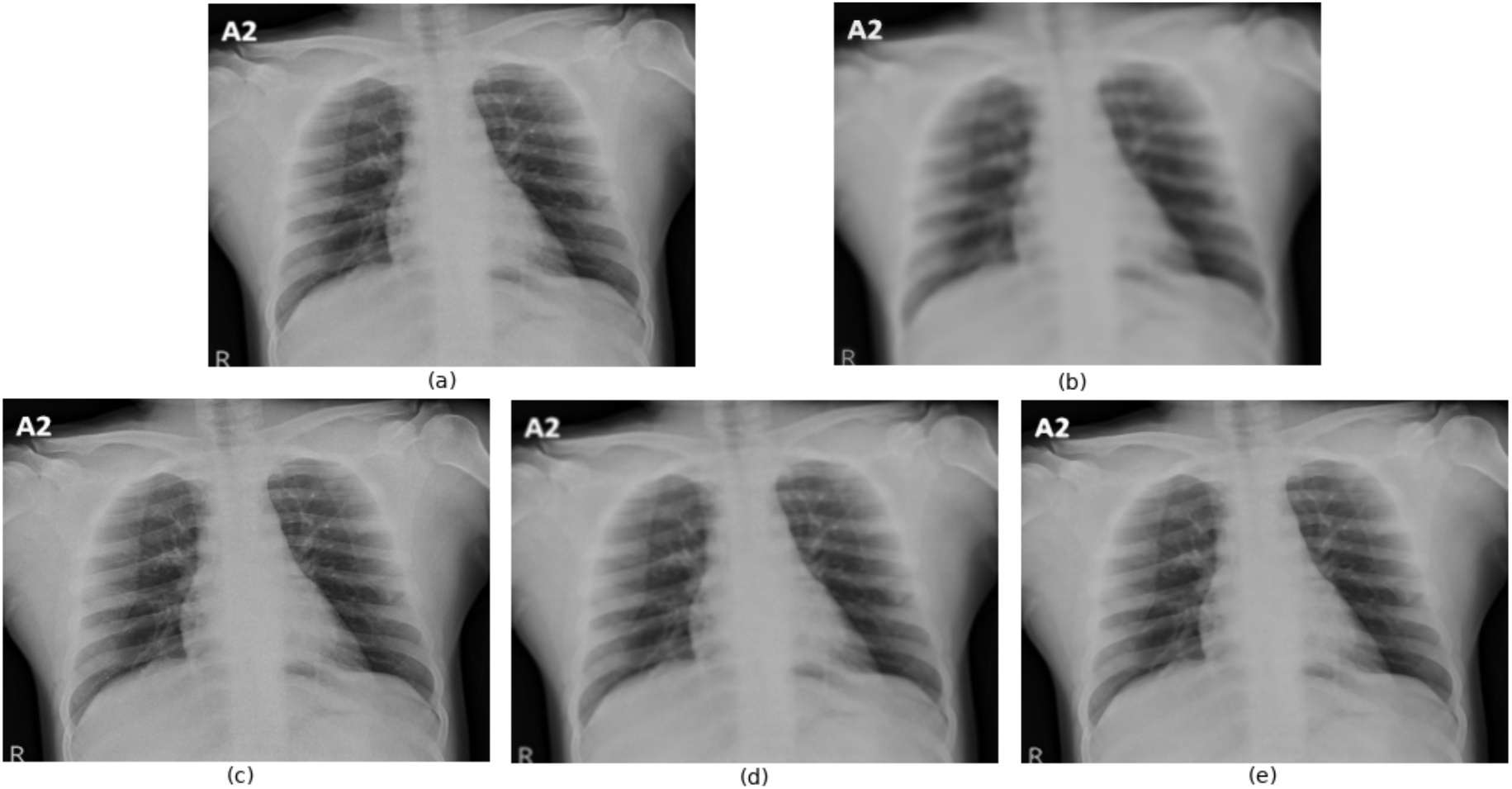
Generated images in augmentation procedure, (a) Original COVID-19 image, (b) Bilateral filtering, (c) Gaussian filtering, (d) Mean filtering and (e) Median filtering.

### Evaluation methods

The properties of the images, the number of images and output classes, and the architectures of proposed or implemented deep networks have a significant effect on the correct detection of COVID-19 in chest X-ray images. Small changes can affect the sensitivity and specificity, as well as recall and precision results produced by a network. Performing training using different folds and the observation of different results also have importance in analysing and assessing the model’s performance. The training and testing were performed by k-fold cross-validation, which divides all the data into a predefined number of folds, *k*, and uses a single fold as a test and the remaining folds for training. The training step is repeated *k* times until all folds have been used for both testing and training. In this study, 8-fold cross-validation was used to evaluate the experiments. A total of 15 images randomly selected from all classes were assigned as the validation set.

During the training phase, augmented data were added to the training folds by matching the training data in the training folds with the corresponding generated data. The process was repeated for each training step, and the considered model was evaluated only by testing the original real-life images. Figure 2 demonstrates the data augmentation and selection procedure in detail.

**Figure 2.**
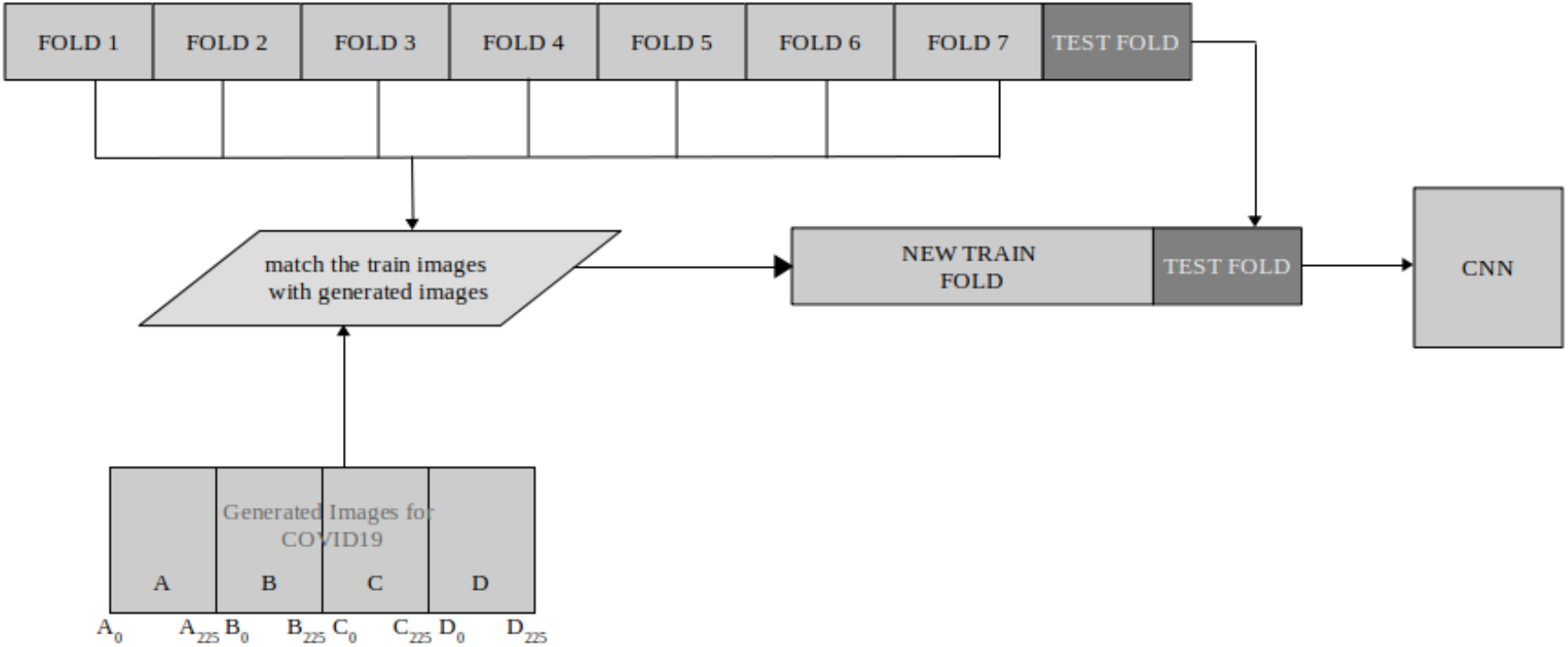
Data augmentation procedure.

The mean accuracy, mean sensitivity and mean specificity were measured; however, the main evaluation for the 2-class-experiments was performed by considering the mean ROC (Receiver Operating Characteristics) Area Under Curve (AUC), which indicates that the model with a higher score is more capable of distinguishing true negatives and true positives.

For the 3-class-experiments, the evaluation was performed by measuring the macro-average F1 score, which considers the recall and precision results produced by the model to determine the general capacity of the model.

## Results

### Results of 2-class-experiments

In the 2-class-experiments, bacterial pneumonia vs COVID-19 and viral pneumonia vs COVID-19 were performed. Two different ConvNet architectures with different input dimensions were considered for the experiments with and without data augmentation procedure, and the evaluation was performed based on the abovementioned criteria.

All ConvNet architectures and image dimensions for the Bacterial Pneumonia and COVID-19 classification produced similar results in terms of mean specificity and mean accuracy; however, fluctuations occurred in mean sensitivity as well as the main criteria, the mean ROC AUC score. CNN1 and CNN2 experiments consisted of three convolutional layers with 128-64-16 filters, and two fully-connected layers with 128 and 32 neurons, respectively. The input image dimensions were 160×120 and 224×224 for the CNN1 and CNN2 experiments, respectively. The ConvNet architecture of CNN3 and CNN4 was created with two convolutional layers (64-32 filters, respectively) and two fully-connected layers with 128 and 8 neurons, respectively. The input dimensions for CNN3 and CNN4 experiments were 160×120 and 80×40, respectively.

The lowest mean sensitivity and mean ROC AUC scores for both experiments with and without data augmentation, were obtained in the CNN2 experiments, which consist the maximum image dimensions considered in this study. The minimum ROC AUC score was 90.67% for all experiments in bacterial pneumonia and COVID-19 classification. The reduced image dimensions in CNN1 and CNN3 experiments without data augmentation achieved ROC AUC scores of 94.00% and 92.86%, respectively. When the generated images were included in the CNN1 and CNN3 experiments, the ROC AUC scores increased to 94.66% and 96.08% respectively; however, the optimal result was achieved in the CNN4 experiment, which had the lowest number of convolutional and fully-connected layers, with the data augmentation procedure. The ROC AUC score obtained in this experiment was 97.32%.

Similar to the bacterial pneumonia and COVID-19 classification experiments, the lowest ROC AUC score for viral pneumonia and COVID-19 classification was obtained in the CNN2 experiment (89.59%). Although increased scores were achieved in the CNN1 and CNN3 experiments, the highest ROC AUC score was again achieved in the CNN4 experiment with the data augmentation procedure (96.80%). Comparison of 2-class-experiments

The obtained results were compared using state-of-the-art pre-trained networks, namely InceptionV3 [18], MobileNet-V2 [19], ResNet50 [20], DenseNet121 [21] and VGG16 [22] by considering both original data without data augmentation procedure, and the generated images.

All pre-trained networks exhibited better performance without the data augmentation procedure, and Densenet121 and Inception-V3 outperformed the other pre-trained networks. Densenet121 produced optimal results in the experiments without data augmentation for both bacterial pneumonia and COVID-19, and viral pneumonia and COVID-19 experiments (96.93% and 95.03%, respectively). However, when the generated images were included, all pre-trained networks failed in convergence and the results showed a sharp fall for all metrics. When all the experiments were considered, the optimal results for bacterial pneumonia and COVID-19, and viral pneumonia and COVID-19 were obtained in the CNN4 experiments with ROC AUC scores of 97.32% and 96.80%, respectively. Table 1 presents all the results obtained in the 2-class-experiments.

**Table 1.**
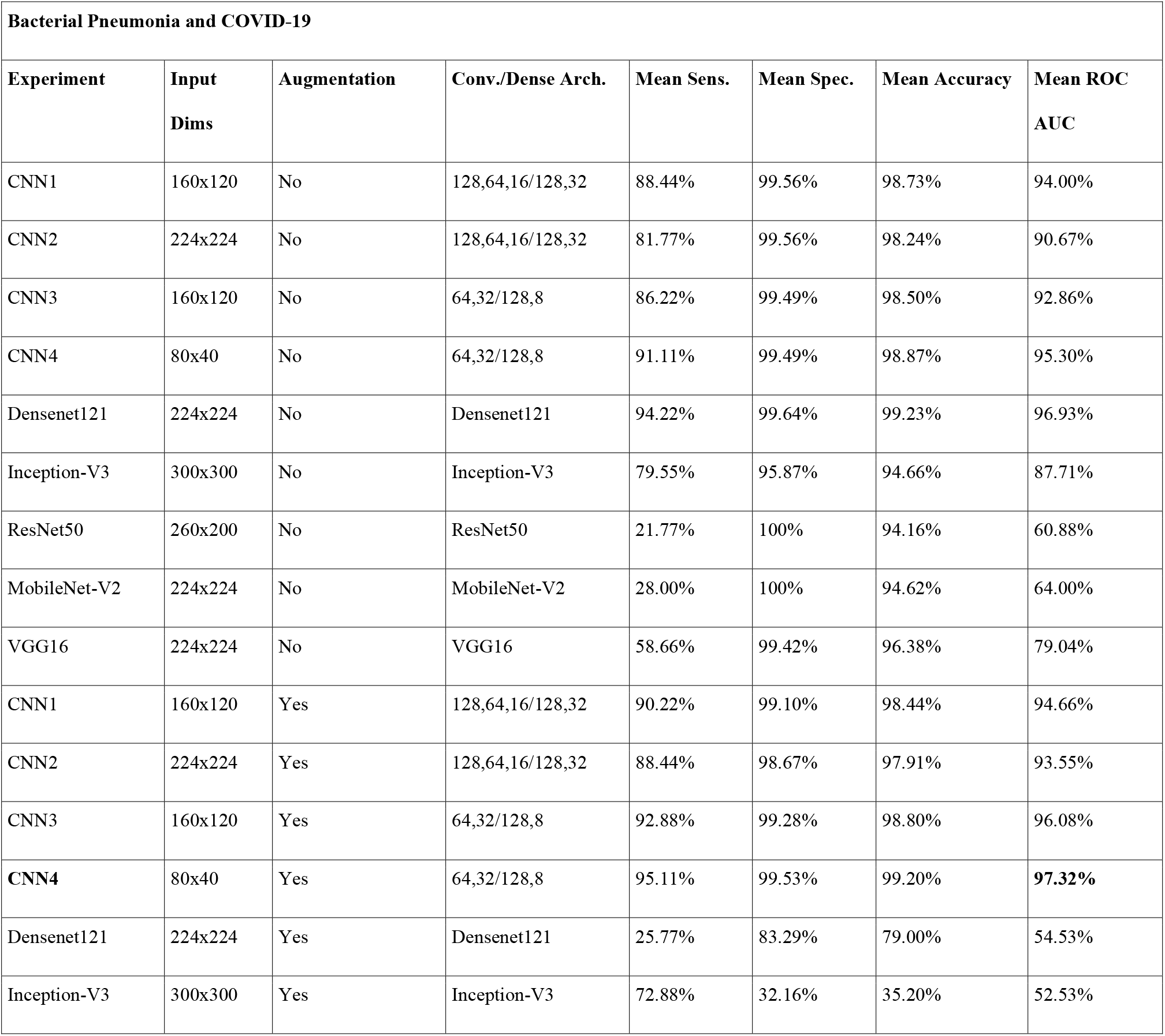
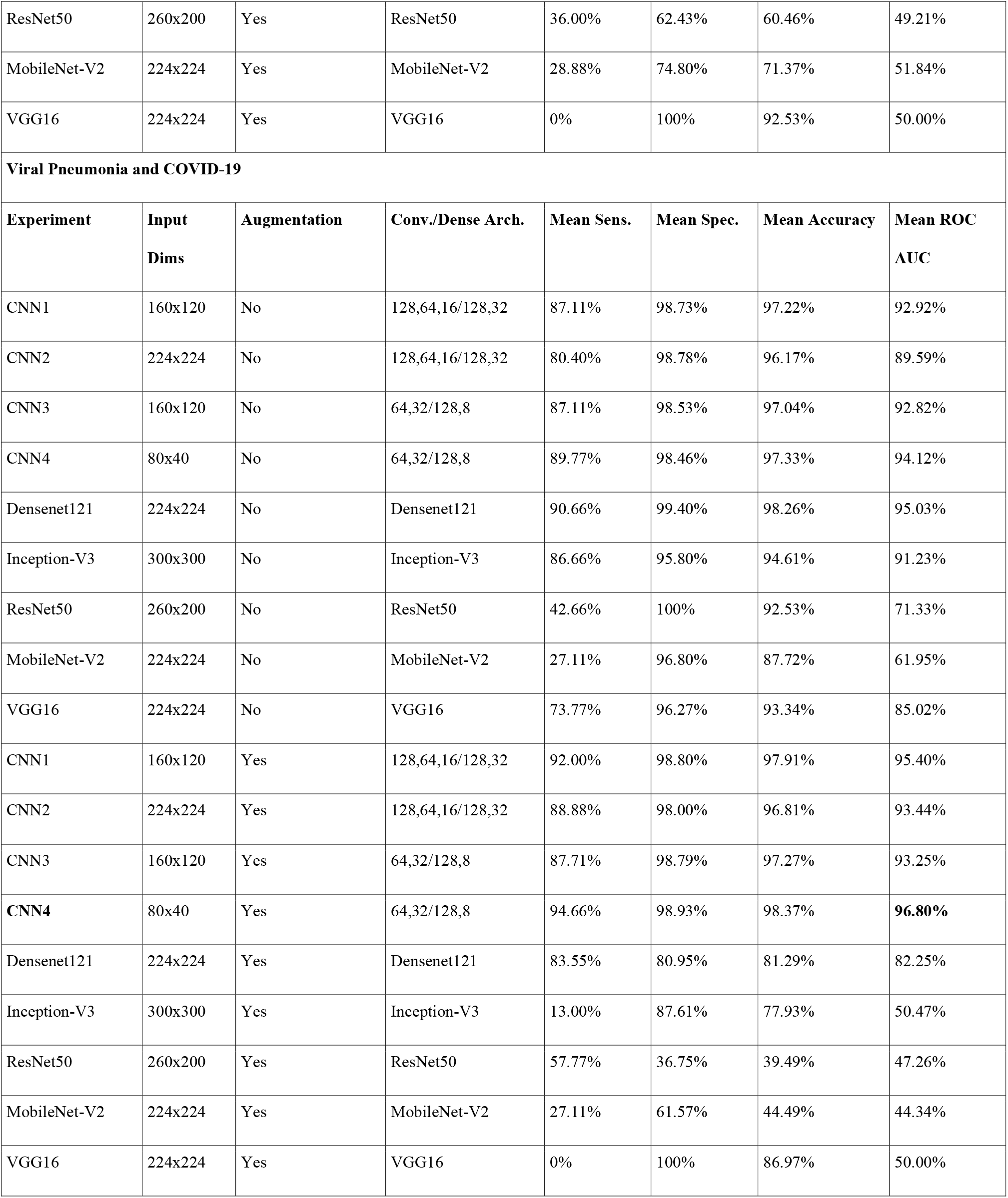
General results for 2-class-experiments

| **Bacterial Pneumonia and COVID-19** |  |  |  |  |  |  |  |
| --- | --- | --- | --- | --- | --- | --- | --- |
| **Experiment** | **Input Dims** | **Augmentation** | **Conv./Dense Arch.** | **Mean Sens.** | **Mean Spec.** | **Mean Accuracy** | **Mean ROC AUC** |
| CNN1 | 160x120 | No | 128,64,16/128,32 | 88.44% | 99.56% | 98.73% | 94.00% |
| CNN2 | 224x224 | No | 128,64,16/128,32 | 81.77% | 99.56% | 98.24% | 90.67% |
| CNN3 | 160x120 | No | 64,32/128,8 | 86.22% | 99.49% | 98.50% | 92.86% |
| CNN4 | 80x40 | No | 64,32/128,8 | 91.11% | 99.49% | 98.87% | 95.30% |
| Densenet121 | 224x224 | No | Densenet121 | 94.22% | 99.64% | 99.23% | 96.93% |
| Inception-V3 | 300x300 | No | Inception-V3 | 79.55% | 95.87% | 94.66% | 87.71% |
| ResNet50 | 260x200 | No | ResNet50 | 21.77% | 100% | 94.16% | 60.88% |
| MobileNet-V2 | 224x224 | No | MobileNet-V2 | 28.00% | 100% | 94.62% | 64.00% |
| VGG16 | 224x224 | No | VGG16 | 58.66% | 99.42% | 96.38% | 79.04% |
| CNN1 | 160x120 | Yes | 128,64,16/128,32 | 90.22% | 99.10% | 98.44% | 94.66% |
| CNN2 | 224x224 | Yes | 128,64,16/128,32 | 88.44% | 98.67% | 97.91% | 93.55% |
| CNN3 | 160x120 | Yes | 64,32/128,8 | 92.88% | 99.28% | 98.80% | 96.08% |
| **CNN4** | 80x40 | Yes | 64,32/128,8 | 95.11% | 99.53% | 99.20% | **97.32%** |
| Densenet121 | 224x224 | Yes | Densenet121 | 25.77% | 83.29% | 79.00% | 54.53% |
| Inception-V3 | 300x300 | Yes | Inception-V3 | 72.88% | 32.16% | 35.20% | 52.53% |
| ResNet50 | 260x200 | Yes | ResNet50 | 36.00% | 62.43% | 60.46% | 49.21% |
| MobileNet-V2 | 224x224 | Yes | MobileNet-V2 | 28.88% | 74.80% | 71.37% | 51.84% |
| VGG16 | 224x224 | Yes | VGG16 | 0% | 100% | 92.53% | 50.00% |
| **Viral Pneumonia and COVID-19** |  |  |  |  |  |  |  |
| **Experiment** | **Input Dims** | **Augmentation** | **Conv./Dense Arch.** | **Mean Sens.** | **Mean Spec.** | **Mean Accuracy** | **Mean ROC AUC** |
| CNN1 | 160x120 | No | 128,64,16/128,32 | 87.11% | 98.73% | 97.22% | 92.92% |
| CNN2 | 224x224 | No | 128,64,16/128,32 | 80.40% | 98.78% | 96.17% | 89.59% |
| CNN3 | 160x120 | No | 64,32/128,8 | 87.11% | 98.53% | 97.04% | 92.82% |
| CNN4 | 80x40 | No | 64,32/128,8 | 89.77% | 98.46% | 97.33% | 94.12% |
| Densenet121 | 224x224 | No | Densenet121 | 90.66% | 99.40% | 98.26% | 95.03% |
| Inception-V3 | 300x300 | No | Inception-V3 | 86.66% | 95.80% | 94.61% | 91.23% |
| ResNet50 | 260x200 | No | ResNet50 | 42.66% | 100% | 92.53% | 71.33% |
| MobileNet-V2 | 224x224 | No | MobileNet-V2 | 27.11% | 96.80% | 87.72% | 61.95% |
| VGG16 | 224x224 | No | VGG16 | 73.77% | 96.27% | 93.34% | 85.02% |
| CNN1 | 160x120 | Yes | 128,64,16/128,32 | 92.00% | 98.80% | 97.91% | 95.40% |
| CNN2 | 224x224 | Yes | 128,64,16/128,32 | 88.88% | 98.00% | 96.81% | 93.44% |
| CNN3 | 160x120 | Yes | 64,32/128,8 | 87.71% | 98.79% | 97.27% | 93.25% |
| **CNN4** | 80x40 | Yes | 64,32/128,8 | 94.66% | 98.93% | 98.37% | **96.80%** |
| Densenet121 | 224x224 | Yes | Densenet121 | 83.55% | 80.95% | 81.29% | 82.25% |
| Inception-V3 | 300x300 | Yes | Inception-V3 | 13.00% | 87.61% | 77.93% | 50.47% |
| ResNet50 | 260x200 | Yes | ResNet50 | 57.77% | 36.75% | 39.49% | 47.26% |
| MobileNet-V2 | 224x224 | Yes | MobileNet-V2 | 27.11% | 61.57% | 44.49% | 44.34% |
| VGG16 | 224x224 | Yes | VGG16 | 0% | 100% | 86.97% | 50.00% |

### Results of 3-class-experiments

The 3-class-experiments were divided into two groups in order to detect COVID-19 among bacterial pneumonia, COVID-19, and healthy group (BCH), and among viral pneumonia, COVID-19, and healthy group (VCH).

By considering the results obtained in the 2-class-experiments, the ConvNet architecture that was used in the CNN4 experiments was considered as the initial stage (Exp_i_) of the 3-class-experiments. The input dimensions were considered as 80×40, and the implementation of ConvNet became deeper for each experiment because of the increased number of images and the output classes, to increase the convergence accuracy.

The ConvNet architecture that was used in the CNN4 experiment achieved macro-average F1 scores of 94.5% and 91.66% of in BCH and VCH experiments without data augmentation procedure, respectively. The macro-average F1 scores obtained with the same architecture and using generated images for BCH and VCH experiments were 94.43% and 92.56%, respectively. The incremental implementation results of the convolutional and fully-connected layers showed that the increment of layers caused small fluctuations in the results, and the optimal macro-average F1 scores were achieved by a ConvNet with four convolutional layers and three fully-connected layers for both BCH and VCH experiments (Exp_l_). In the BCH experiments, macro-average F1 scores of 95.79% and 94.67% were achieved for the experiments with data augmentation and without data augmentation, respectively. In the VCH experiments, macro-average F1 scores of 94.59% and 93.51% of were obtained with and without generated images, respectively.

#### Comparison of 3-class-experiments

Similar to the previous comparisons, the obtained results were compared by state-of-the-art pre-trained networks; however, only the InceptionV3 [18] and DenseNet121 [21] were included because of the high ROC AUC scored obtained in the 2-class-experiments without data augmentation procedure.

The considered pre-trained networks produced similar but not optimal results when original images were used during training; however, they were not able to detect COVID-19 effectively, using augmented data. Table 2 shows the macro-average F1 scores obtained for the pre-trained networks and two ConvNets as initial (Exp_i_) and the final (Exp_l_) architecture which produced the optimal results.

**Table 2.** Results for 3-class-experiments

| **BCH Experiments** |  |  |  |
| --- | --- | --- | --- |
| **Experiment** | **Augmentation** | **Conv./Dense Arch.** | **Macro-average F1 score** |
| Exp<sub>i</sub> | NO | 64,32 / 128,8 | 94.50% |
| Exp<sub>f</sub> | NO | 128,128,64,32 / 128,64,128 | 94.67% |
| Densenet121 | NO | Densenet121 | 94.10% |
| Inception-V3 | NO | Inception-V3 | 92.93% |
| Exp<sub>i</sub> | YES | 64,32 / 128,8 | 94.43% |
| Exp<sub>f</sub> | YES | 128,128,64,32 / 128,64,128 | **95.79%** |
| Densenet121 | YES | Densenet121 | 16.66% |
| Inception-V3 | YES | Inception-V3 | 22.14% |
| **VCH Experiments** |  |  |  |
| Exp<sub>i</sub> | NO | 64,32 / 128,8 | 91.66% |
| Exp<sub>f</sub> | NO | 128,128,64,32 / 128,64,128 | 93.51% |
| Densenet121 | NO | Densenet121 | 92.33% |
| Inception-V3 | NO | Inception-V3 | 88.89% |
| Exp<sub>i</sub> | YES | 64,32 / 128,8 | 92.56% |
| Exp<sub>f</sub> | YES | 128,128,64,32 / 128,64,128 | **94.59%** |
| Densenet121 | YES | Densenet121 | 34.33% |
| Inception-V3 | YES | Inception-V3 | 28.45% |

## Discussion

Analysis of the experiments and the obtained results should be performed according to the input and output properties of ConvNet. In the results of the 2-class-experiments, which were performed to detect two types of diseases, namely COVID-19 and viral pneumonia or bacterial pneumonia, the decrement of input dimensions, and the use of a deep but lighter ConvNet architecture produced more accurate results than other considered architectures. However, in the 3-class-experiments, which had three outputs in the ConvNet architecture, the obtained results suggested that a deeper architecture should be used for more efficient convergence and test results.

The data augmentation procedure was applied for all experiments and increased test scores were observed for all experiments in which the generated data were considered. The increase in scores varied between 1.5% and 4.0% for all experiments.

The results produced by the pre-trained networks, DenseNet121 and Inception-V3, without using generated data, were stable and similar to the experiments performed in this study; however, when the augmented data were added to the training phase of the pre-trained networks, over-fitting occurred during the convergence and the test results suddenly decreased. The results obtained in the two different categories demonstrates that filtering approaches can be used in data augmentation procedures to increase the accuracy of the model; however, the fixed layered ConvNet cannot be effectively used for the detection of COVID-19 and other pneumonia types.

Researchers have performed several experiments by implementing different ConvNet architectures and pre-trained networks to detect different variations of COVID-19, healthy and other pneumonia types using X-ray chest images. Different image databases with different numbers of images for each disease were considered, and the evaluation metrics varied in these studies. Therefore, although performing a head-to-head comparison is not possible, according to the results obtained in this study and recent studies, it is possible to obtain a good idea.

In a study by Zhang et al. [23], COVID-19 and non-COVID-19 images were trained and the obtained results were evaluated by ROC AUC score. They concluded that an ROC AUC score of 95.2% of was achieved in their study with their proposed model. Apostolopoulos & Mpesiana [24] achieved an accuracy of 93.48% using a pre-trained network VGG19 by considering COVID-19, healthy and pneumonia images. Hemdan et al. [25] trained several pre-trained networks for COVID-19 detection in their proposed COVIDX-Net framework. They used F1-scores for evaluating the model and it was concluded that the VGG19 and DenseNet201 models could effectively be applied for that purpose. Sethy and Behera [26] implemented ResNet50 and SVM to detect COVID-19 and an accuracy of 95.4% was achieved in their study. Table 3 presents the results obtained in both recent researches and this study in detail.

**Table 3.** The results obtained in this study and recent studies. B: Bacterial, C: COVID-19, H: Healthy, P: Pneumonia, V: Viral.

| Classification | Optimal Method | Result (%) | Reference |
| --- | --- | --- | --- |
| C / P | Proposed Model | 95.18 AUC | Zhang et al. [23] |
| C / H | VGG19 / DenseNet201 | 91.00 F1 score | Hemdan et al. [25] |
| C / H | ResNet50 + SVM | 95.52 F1 score | Sethy and Behera [26] |
| C / BP / H | VGG19 | 93.48 Accuracy | Apostolopoulos & Mpesiana [24] |
| C / P / H | MobileNet v2 | 94.72 Accuracy | Apostolopoulos & Mpesiana [24] |
| C / P / H | DarkNet | 87.02 Accuracy | Ozturk et al. [12] |
| C / BP+VP / H | Bayes-SqueezeNet-based | 98.26 Accuracy | Ucar & Korkmaz [13] |
| C / BP | Augmented Lighter ConvNet | 97.32 Mean ROC AUC | This Study |
| C / VP | Augmented Lighter ConvNet | 96.80 Mean ROC AUC | This Study |
| C / BP / H | Augmented Deeper ConvNet | 95.79 Macro-average F1 score | This Study |
| C / VP / H | Augmented Deeper ConvNet | 94.59 Macro-average F1 score | This Study |

The early diagnosis of COVID-19 has become a research focus among radiologist and clinicians across the world. Image clarification using AI techniques can help in the early detection of the disease. In this study, different ConvNets for the diagnosis of COVID-19 were proposed to achieve more accurate results compared to the results in the existing literature for the diagnosis of the disease. The proposed method was able to distinguish COVID-19 images among non-COVID-19 images, namely bacterial and viral pneumonia as well as normal X-ray images. The use of different augmentation techniques that produce similar images with different characteristics can help the convergence of the model and improve the obtained scores. In addition, different deep architectures should be analyzed in further studies to achieve optimal results.

## Data Availability

Data used in this study is available in the main text.

https://github.com/ieee8023/covid-chestxray-dataset

https://doi.org/10.17632/rscbjbr9sj.3

## Notes

### Competing Interest Statement

The authors have declared no competing interest.

### Funding Statement

The authors received no specific funding for this work.

### Author Declarations

The project built for creating this dataset was approved by the University of Montreal's Ethics Committee #CERSES-20-058-D. Informed consent was obtained for all subjects, and the study was approved by the relevant institutional review board at each data acquisition site. All methods were performed in accordance with the relevant guidelines and regulations. All chest X-ray imaging was performed as part of patients' routine clinical care. Institutional Review Board (IRB)/Ethics Committee approvals were obtained. The work was conducted in a manner compliant with the United States Health Insurance Portability and Accountability Act (HIPAA) and was adherent to the tenets of the Declaration of Helsinki.

